# CD8+ tissue-resident memory T cells triggered the erosion of oral lichen planus by the cytokine network

**DOI:** 10.1101/2022.10.18.22281149

**Authors:** Maofeng Qing, Qianhui Shang, Dan Yang, Jiakuan Peng, Jiaxin Deng, Lu Jiang, Jing Li, Yu Zhou, Hao Xu, Qianming Chen

## Abstract

CD8^+^ tissue-resident memory T (CD8^+^ Trm) cells play key roles in many immune-inflammation-related diseases. However, their characteristics in the pathological process of oral lichen planus (OLP) are unclear. Therefore, we investigated the function of CD8^+^ Trm cells in the process of OLP. Single-cell RNA sequencing profiling and spatial transcriptomics revealed that compared with non-erosive OLP, CD8^+^ Trm cells, which were mainly distributed in the lamina propria close to the basement membrane, were increased and functionally more active by secreting multiple cytokines in patients with erosive oral lichen planus (EOLP), including IFN-γ, TNF-α, and IL17. And our clinical cohort of 1-year follow-up was also supported the above results in RNA level and protein level. In summary, this study provided a novel molecular mechanism for triggering OLP erosion by CD8^+^ Trm cells to secrete multiple cytokines, and new insight into the pathological development of OLP.

## Introduction

Oral lichen planus (OLP) is a chronic inflammatory disease on the oral mucosa of unknown etiology mediated by T cells with a 1.01% global prevalence rate (Gonzalez-Moles et al., 2021; Jiang et al., 2022b). Its clinical manifestations are varied and chronic non-healing (Radwan-Oczko, 2013). Especially, recurrent erosion or ulcers were mostly accompanied by pain and discomfort, which worsen when eating or speaking, and adversely affect the physical and mental health of patients (Rotaru et al., 2020).

The pathological feature of OLP is characterized by dense infiltration of T cells in the lamina propria, disruption of the basement membrane, and degeneration of basal keratinocytes (Xu et al., 2022). Previous studies have found that there are immune function disorders in the local and systemic lesions of OLP patients, and the infiltration of CD8^+^ T cells in the OLP lamina propria near the basement membrane (Neppelberg et al., 2001). CD8^+^ T cells are related to the liquefaction of basal cells, and it is believed that the immune microenvironment of OLP lesions has changed, especially in CD8^+^ T cells, which produce a large number of cytokines and chemokines, such as IFN-γ, TNF-α, IL-1α, and IL-17, forming a complex cytokine network (Firth et al., 2015; Ke et al., 2017; Piccinni et al., 2014; Shaker and Hassan, 2012; Viguier et al., 2015).

Tissue-resident memory T (Trm) cells are a recently described population of terminally differentiated T cells, which are confirmed to have an important relationship with local immunity (Mueller et al., 2014). It was found that Trm cells are mainly present in various barrier tissues, and CD8^+^ Trm cells can persist locally for a long time in the absence of relevant antigens (Szabo et al., 2019). Recent work has found that CD8^+^ Trm cells play important roles in the occurrence and development of many chronic inflammatory diseases such as psoriasis and vitiligo (Eberle et al., 2016; Richmond et al., 2018; Watanabe, 2019).

In contrast, the presence, distribution, and role of CD8^+^ Trm cells in OLP are unclear. This work sought to investigate the presence, spatial position, and function of CD8^+^ Trm cells in OLP, especially in erosive OLP. And this study examined the heterogeneity of CD8^+^ Trm cells in different clinical types and deeply explored the molecular mechanism of the erosion of OLP.

## Results

### Single-cell RNA sequencing revealed the cell composition of OLP with different clinical subtypes

To investigate the cellular composition and comprehensive transcriptional effects of OLP, we performed single-cell RNA sequencing (scRNA-seq) of non-erosive OLP (NEOLP, n = 3) and erosive OLP (EOLP, n = 1). The final dataset comprised 31,330 cells, with an average of 1470 genes per cell. Visualization using uniform manifold approximation and projection (UMAP) revealed 28 distinct cell clusters (Figure 1A) that were annotated as 10 major cell types (Figure 1B). Although there was no significant difference in T cell proportion between NEOLP and EOLP, T cell was still the major cell proportion in EOLP Surprisingly, although T cells, the hallmark cells of OLP, constituted the highest proportion of all samples, the proportion of cells in EOLP was the lowest at 65.36%, while the proportion of T cells in the NEOLP-3 case is the highest at 82.88%. The cell composition analysis also revealed an increase in the proportion of B cells and mast cells in EOLP (Figures 1C and 1D).

**Fig. 1.**
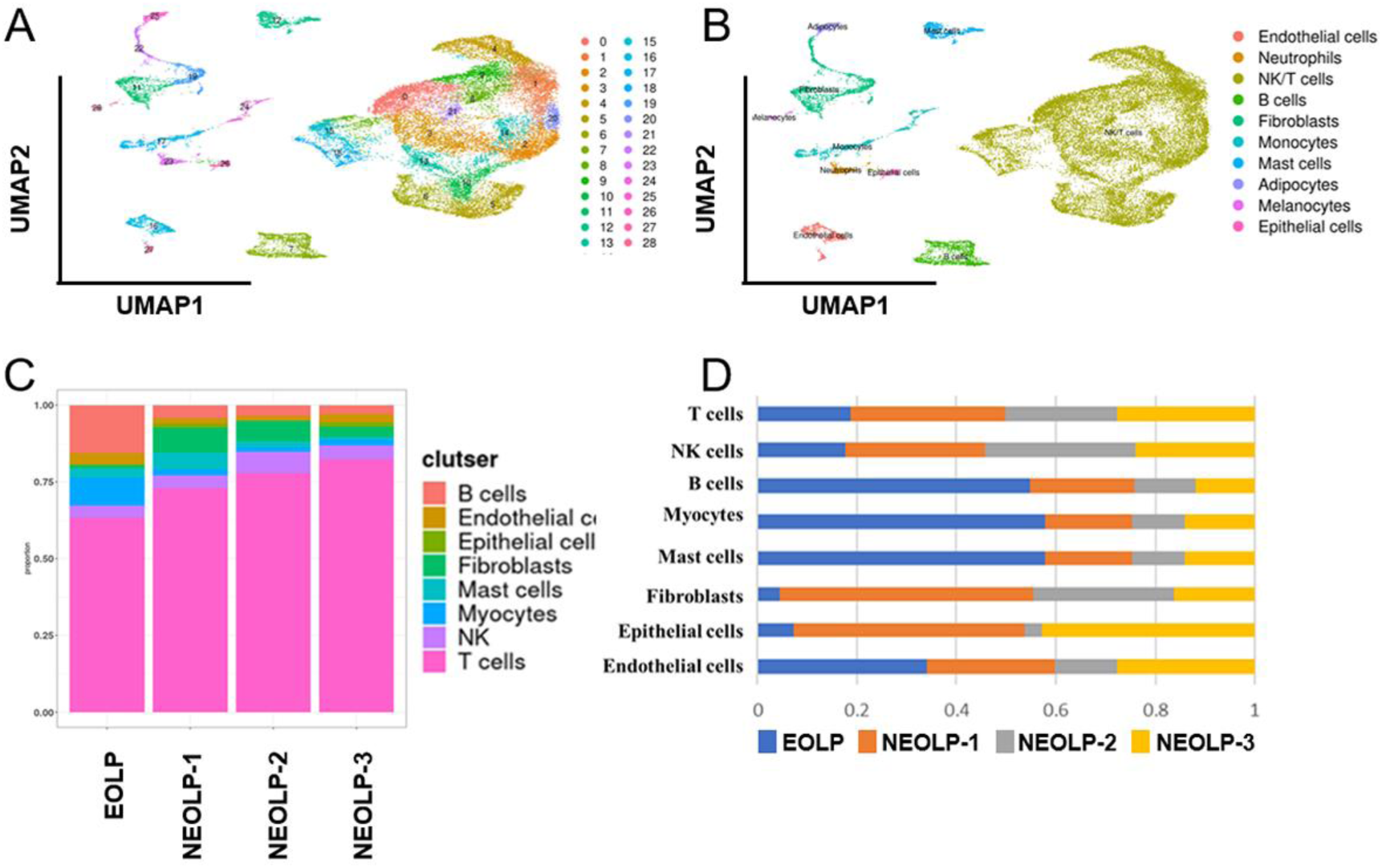
scRNA-seq capture the cellular in patients with NEOLP and EOLP. (B) UMAP plot of 31,330 cells colored by cell type. (C) Bar plot showing the percentage of cell subsets in different samples. (D) The proportion of each sample in different cell subsets.

### The cell proportion of CD8^+^ Trm cells was increased in EOLP

The typical pathological manifestation of OLP is band infiltration of T cells in the lamina propria. At present, most views believe that OLP is an immune-related disease mediated by T cells (Feldmeyer et al., 2020). So, our next further analyzed the T cell population.

Since NK and T cells are also derived from lymphoid progenitor cells, NK and T cells are developmentally closer and mature NK cells express CD3 (a T cell signature gene) after activation, while the cytotoxic response of mature NK cells is similar to that of CTL (Abel et al., 2018). Therefore, in the initial study, T cells and NK cells were included together to form separate NK and T cell populations, and the NK/T population constituted the majority of cells sequenced in the OLP in the study data. Subsequently, by further clustering, the genes of the NK/T cell population, the NK/T cell population was divided into a total of 11 cell subgroups (Figures S1A, and S1B). After further annotation of the NK/T cell subsets with marker genes, six large cell subpopulations were described: NK cells, CD8^+^ T cells, CD4^+^ T cells, CD4^+^ CD8^+^ T cells, CD8^+^ Trm cells, CD4^+^ CD8^+^ Trm cells (Figures 2A and 2B).

**Fig. 2.**
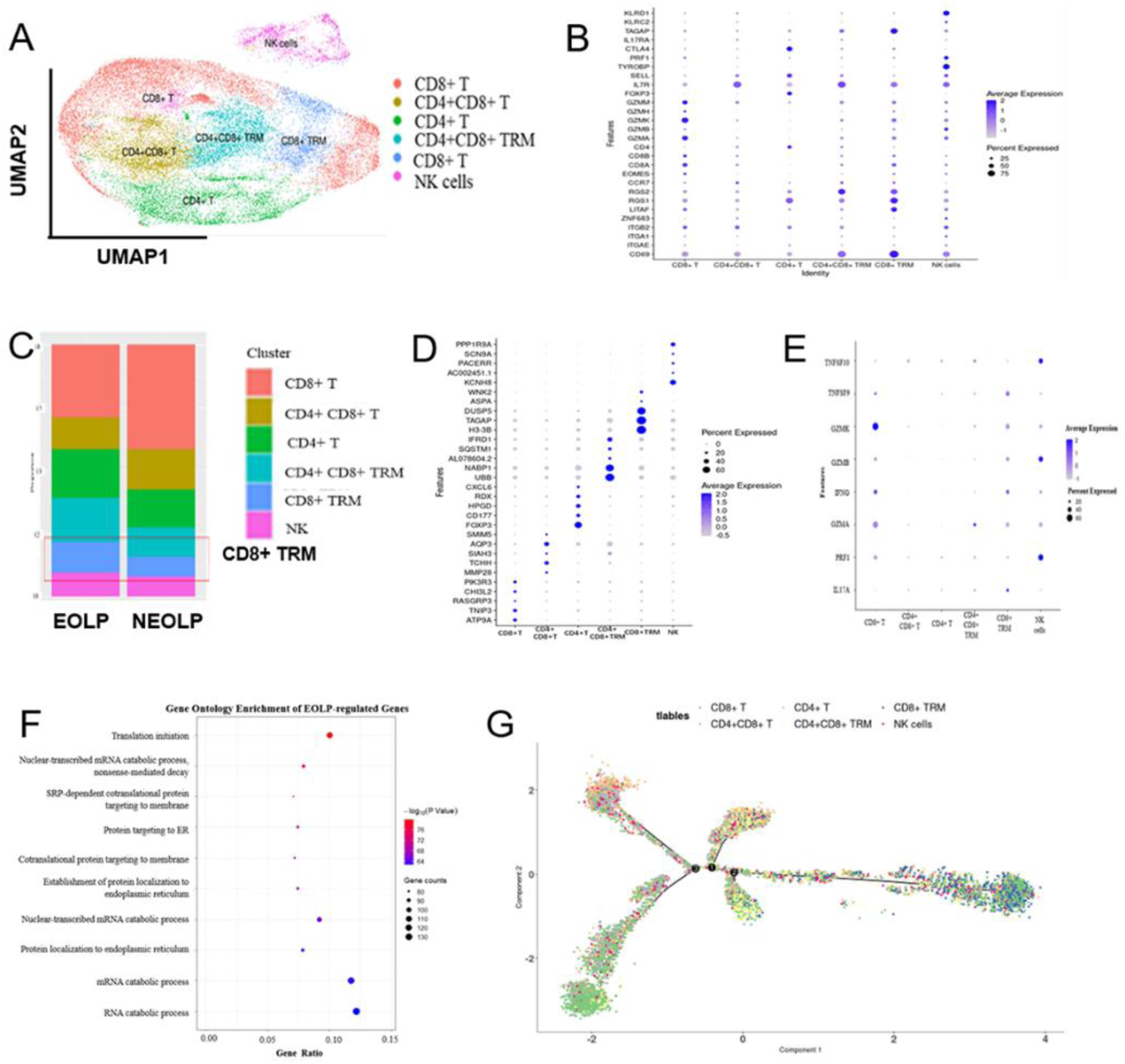
CD8+ TRM in OLP patients have different transcriptomic landscapes in different clinical presentations. (A) UMAP plot of NK/T cells colored by subtype. (B) Dot plot of marker genes in NK/T cells subsets. (C) Proportion of NK/T cell subsets in OLP with different clinical manifestations. (D) Dot plot of significant differentially expressed genes (DEGs) in NK/T cells subsets. (E) Dot plot of differential expression of inflammatory cytokine product genes in NK/T cell subsets. (F) Dot plot of GO enrichment analysis of DEGs in CD8+ TRM subsets between EOLP and NEOLP. (G) Pseudotime trajectory of NK/T cells in OLP.

Significant changes were observed in the proportion of T cell populations in patients with different clinical types of OLP, with fewer CD8^+^ T cells, CD4^+^ CD8^+^ T cells in EOLP than in NEOLP, while CD4^+^ T cells, CD8^+^ Trm cells, CD4^+^ CD8^+^ Trm cells increased in EOLP, and the proportion of NK cells in the two was basically the same (Figure 2C).

### The immune function of CD8^+^ Trm cells was activated in EOLP

To further explore the function of T cells, this study exhibited that in the CD8^+^ Trm cells subset, the T cell activation gene TAGAP was significantly up-regulated, while in the CD4^+^ T cell subset, the gene FOXP3, which regulates the development and regulation of Treg cells (Kane et al., 2013), was significantly activated, accompanied by high expression of some cell exhaustion genes (e.g., CD177, HPGD, and RDX) (Figures 2D and S1C) (Kane et al., 2013).

When pro-inflammatory cytokines like tumor necrosis factor (TNF), interferon-gamma (IFN-γ), granzyme, perforin, and IL-17 were compared between each subset, it was discovered that CD8+ Trm cells were the cells most crucial for producing IL-17. Additionally, it exhibited a significant capacity to create TNF and IFN-γ, which may have contributed to the worsening of the clinical manifestations of OLP (Figure 2E). Subsequently, we performed differential expression analysis between CD8^+^ Trm cells in EOLP versus NEOLP and found regulatory genes of multiple pro-inflammatory cytokines were significantly upregulated in EOLP (e.g., RORA, IL32, and CCL20) (Figure 2F) (Ito et al., 2011; Kim et al., 2005; Lluis et al., 2014). Compared with NEOLP, the transcription genes of the CD8 ^+^ Trm subset, Runx-1 and IL23A, were significantly increased in EOLP, and Runx-1 can increase the expression of IL2 and IFN-γ (Ono et al., 2007). After IL23A binds to the receptor, it has the function of activating memory T cells, which can promote the secretion of IFN-γ (Piskin et al., 2006). IL-17/IL-23 Axis plays an important role in many autoimmune diseases (Dragasevic et al., 2018). It has been proved that the IL-17/IL-23 Axis plays an important role in the pathogenesis of OLP by our research group (Lu et al., 2014). GO enrichment analysis indicated that CD8^+^ Trm DEGs in EOLP between NEOLP were more significantly enriched in pathways such as translation initiation (Fig 2J). It shows that CD8^+^ Trm subsets in EOLP have different states and expressions compared with NEOLP subsets. The pseudotime analysis also suggests that CD8^+^ Trm cells is one of the terminal states of T cell differentiation and development in OLP (Figure 2G), which may have a profound relationship with their clinical manifestations.

### Spatial transcriptomics revealed CD8^+^ Trm cells were adjacent to the epithelium of OLP and its products may induce epithelial erosion and promote the development of disease

Cell types, relative positions among cells, and the levels of gene expression of cell populations together determine their function in biological tissues. To investigate the spatial heterogeneity between normal oral mucosa and OLP, we performed the Spatial Transcriptomics (ST) analysis of normal oral mucosa (n = 2), NEOLP (n = 3), and EOLP (n = 1).

The tissues in this study covered spots ranging from the lowest 296 spots in the NEOLP-1 to the highest 965 spots in the NEOLP-3, while the per-sample capture factors ranged from 2869 to 3884 (Table S1). We first used keratin to characterize the basal layer of the epithelium. It can be seen that in the normal oral mucosa and NEOLP, the epithelium was intact, while in the NEOLP, some samples had atrophied and thinned epithelium, while some samples had hyperplasia and thickening. In EOLP, it can be seen that the epithelium on the left side of the tissue is intact, while the epithelium on the right side is missing (Figure 3A).

**Fig. 3.**
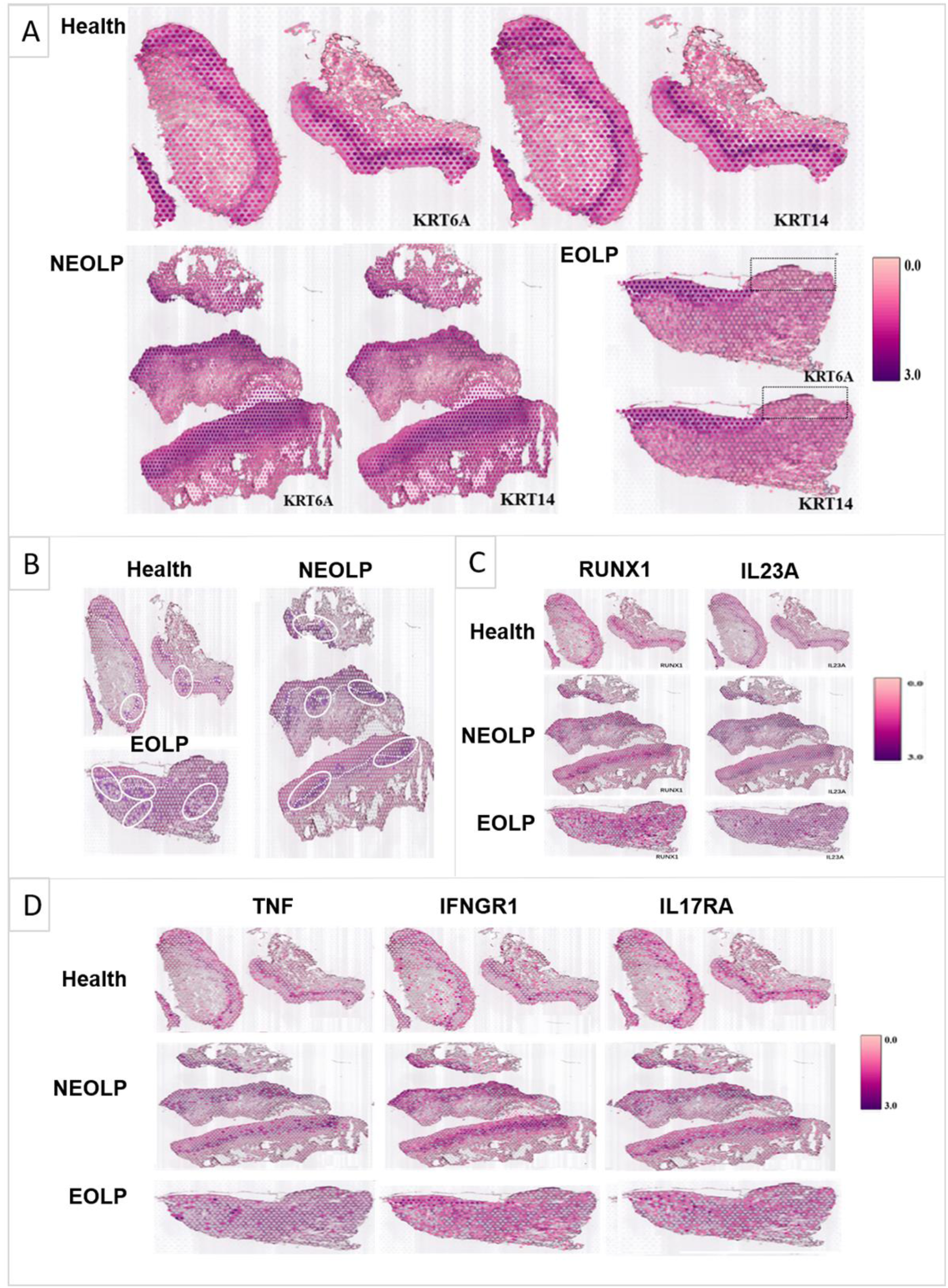
CD8+ TRM in OLP patients have different transcriptomic landscapes in different clinical presentations. (A) Marked the epithelial layer area of each group of samples. B) CD8+ TRM distribution area. The dark purple spots are the CD8+ TRM marker gene expression spots; the area in the white circle is the CD8+ TRM distribution area; the white dotted line is the basement membrane. (C) RUNX1/IL23A distribution area. (D) TNF/IFNGR1/IL17A/IL17RA distribution area.

After co-localization with the marker gene of CD8^+^ Trm cells, it was found that CD8^+^ Trm cells were more distributed in EOLP than NEOLP, and more distributed in NEOLP than normal oral mucosa. And it can be observed that whether in normal oral mucosa, NEOLP or EOLP, CD8^+^ Trm cells are mostly distributed adjacent to the epithelium, while in EOLP tissues, CD8^+^ Trm cells in the areas where the epithelium is lost are correspondingly reduced, and there are CD8^+^ Trm cells in the deeper lamina propria of the corresponding tissues (Figure 3B). It indicated that CD8^+^ Trm cells may be closely related to OLP epithelium. Consistent with the results of scRNA-seq, the expression of RUNX1 and IL23A was more pronounced in the epithelial deletion region in erosive OLP, which may be related to the clinical manifestations of OLP.

In OLP tissue, the expression of TNF, IL17A/IL17RA, IFNGR1, etc. was higher than in normal oral mucosa. And the signals of the effector receptor IFNGR1 of IFN and the effector receptor IL17RA of IL17 were significantly enhanced in EOLP compared with NEOLP.

### Cohort studies confirmed the core genes of CD8^+^ Trm cells were closely associated with the erosion and process of OLP

To objectively illustrate the molecular mechanism of OLP erosion, we established a clinical cohort with 40 participants, which were confirmed by clinical manifestation and pathological diagnosis. The basic information of the clinical cohort was shown in Table 1. This study included 40 OLP patients, there were 15 females and 12 males in the NEOLP group and 8 females and 5 males in the EOLP group. The mean age of the NEOLP group was 30.07±11.34 years and that of the EOLP group was 45.08± 12.86 years. There were 7 smoking patients and 9 drinking patients in NEOLP and 2 smoking patients and 3 drinking patients in EOLP. The mean VAS score of the NEOLP group was 1.41± 0.89 and that of the EOLP group was 2.46± 1.391. The mean course of disease was 14.85± 27.932 months in the NEOLP group and 14.85± 27.932 months in the EOLP group. There was no significant difference in clinical information between the two groups (Table 1).

**Table 1.** Analysis of clinical variables in different clinical types of OLP.

| Factors | NEOLP | EOLP | $t/c^2$ | $p$ |
| --- | --- | --- | --- | --- |
| Gender |  |  | 0.129 | 0.72 |
| Female | 15 | 8 |  |  |
| Male | 12 | 5 |  |  |
| Age(years) | 39.07±11.34 | 45.08±12.86 | -1.502 | 0.348 |
| Smoking |  |  | 0.559 | 0.455 |
| Yes | 7 | 2 |  |  |
| No | 20 | 11 |  |  |
| Drinking |  |  | 0.44 | 0.507 |
| Yes | 9 | 3 |  |  |
| No | 18 | 10 |  |  |
| VAS scoring | 1.41±0.89 | 2.46±1.391 | -2.91 | 0.341 |
| Course of disease (months) | 14.85±27.932 | 23.38±43.065 | -0.755 | 0.12 |

Through differential gene analysis, it was found that in NEOLP and EOLP, the expression of multiple core factors and effector cytokines or their receptor transcription factors of CD8^+^ Trm cells, including ITGA1, LITAF, SKIL, etc., and related immune factor receptors such as TNFRSF6B, IL17RA, IFNAR1, etc., were significantly increased in the EOLP than in the NEOLP (Figures S2A and S2B; Table 2).

**Table 2.**
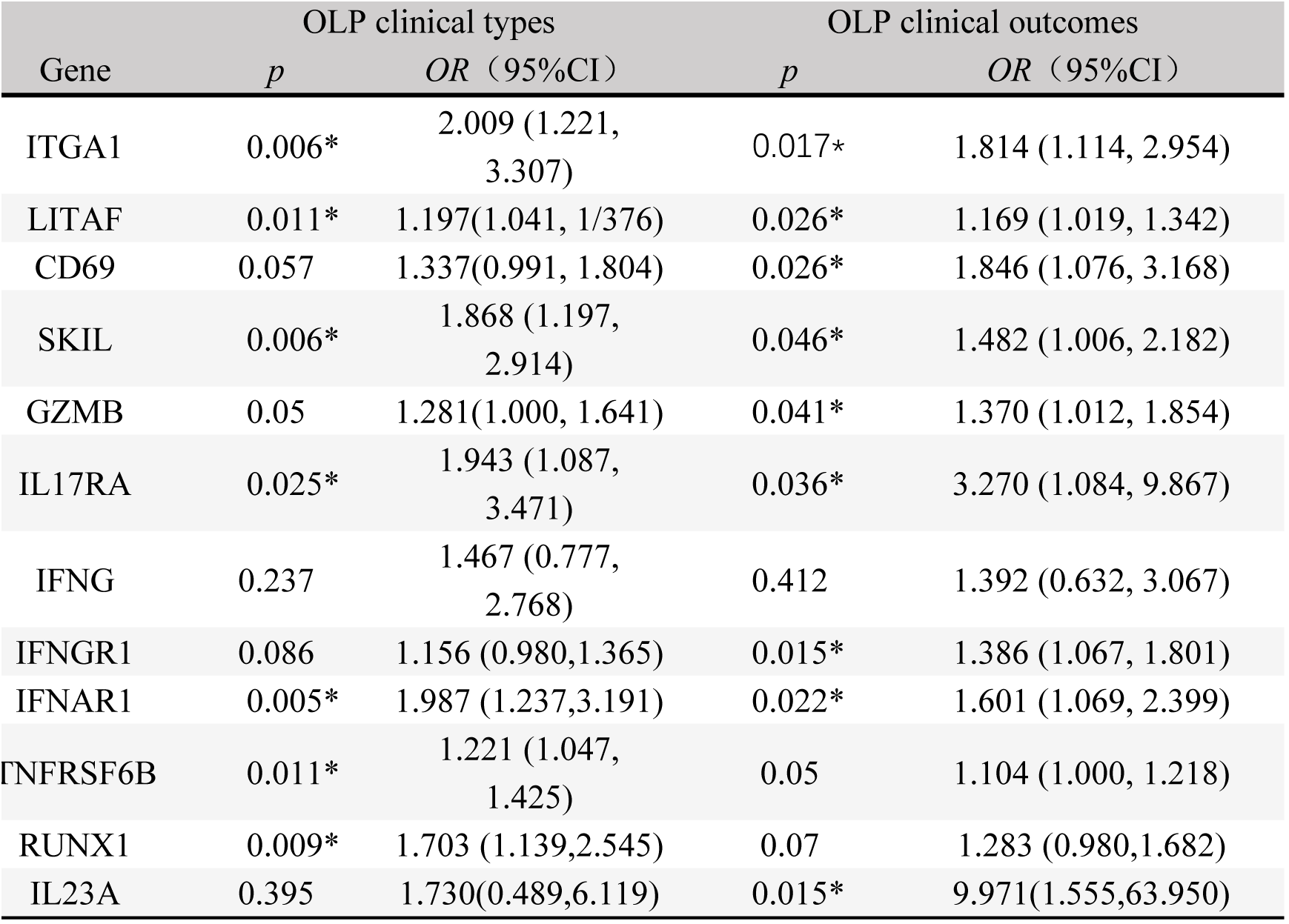
Logistic regression analysis of CD8+ Trm core transcriptome in different OLP clinical types and clinical outcomes.

| Gene | OLP clinical types |  | OLP clinical outcomes |  |
| --- | --- | --- | --- | --- |
| | $p$ | OR (95%CI) | $p$ | OR (95%CI) |
| ITGA1 | 0.006* | 2.009 (1.221, 3.307) | 0.017* | 1.814 (1.114, 2.954) |
| LITAF | 0.011* | 1.197(1.041, 1/376) | 0.026* | 1.169 (1.019, 1.342) |
| CD69 | 0.057 | 1.337(0.991, 1.804) | 0.026* | 1.846 (1.076, 3.168) |
| SKIL | 0.006* | 1.868 (1.197, 2.914) | 0.046* | 1.482 (1.006, 2.182) |
| GZMB | 0.05 | 1.281(1.000, 1.641) | 0.041* | 1.370 (1.012, 1.854) |
| IL17RA | 0.025* | 1.943 (1.087, 3.471) | 0.036* | 3.270 (1.084, 9.867) |
| IFNG | 0.237 | 1.467 (0.777, 2.768) | 0.412 | 1.392 (0.632, 3.067) |
| IFNGR1 | 0.086 | 1.156 (0.980,1.365) | 0.015* | 1.386 (1.067, 1.801) |
| IFNAR1 | 0.005* | 1.987 (1.237,3.191) | 0.022* | 1.601 (1.069, 2.399) |
| TNFRSF6B | 0.011* | 1.221 (1.047, 1.425) | 0.05 | 1.104 (1.000, 1.218) |
| RUNX1 | 0.009* | 1.703 (1.139,2.545) | 0.07 | 1.283 (0.980,1.682) |
| IL23A | 0.395 | 1.730(0.489,6.119) | 0.015* | 9.971(1.555,63.950) |

On the other hand, according to the clinical manifestation in the 1-year follow-up, the clinical cohort was classified into two groups, the recurrent erosion (RE) group (the interval between erosions <3 months) and the persistent non-erosion (PNE) group (≥3 months without any form of erosion). In addition to the above clinical factors being included in the two groups for statistical comparison, the diagnosis (NEOLP or EOLP) and medication (divided into three groups: local glucocorticoids, local glucocorticoids ^+^ immunosuppressant, and other drugs) were analyzed (Table 3), in which glucocorticoids were all used for local and external use. Differences in clinical information other than diagnosis were still not statistically significant for the clinical outcome of OLP. The results of multiple core factors and effector cytokines of CD8^+^ Trm cells in the recurrent erosion group were consistent with the results of EOLP, which confirm the consistency and accuracy of the results of EOLP with less time influence (Figures S2C and S2D).

**Table 3.**
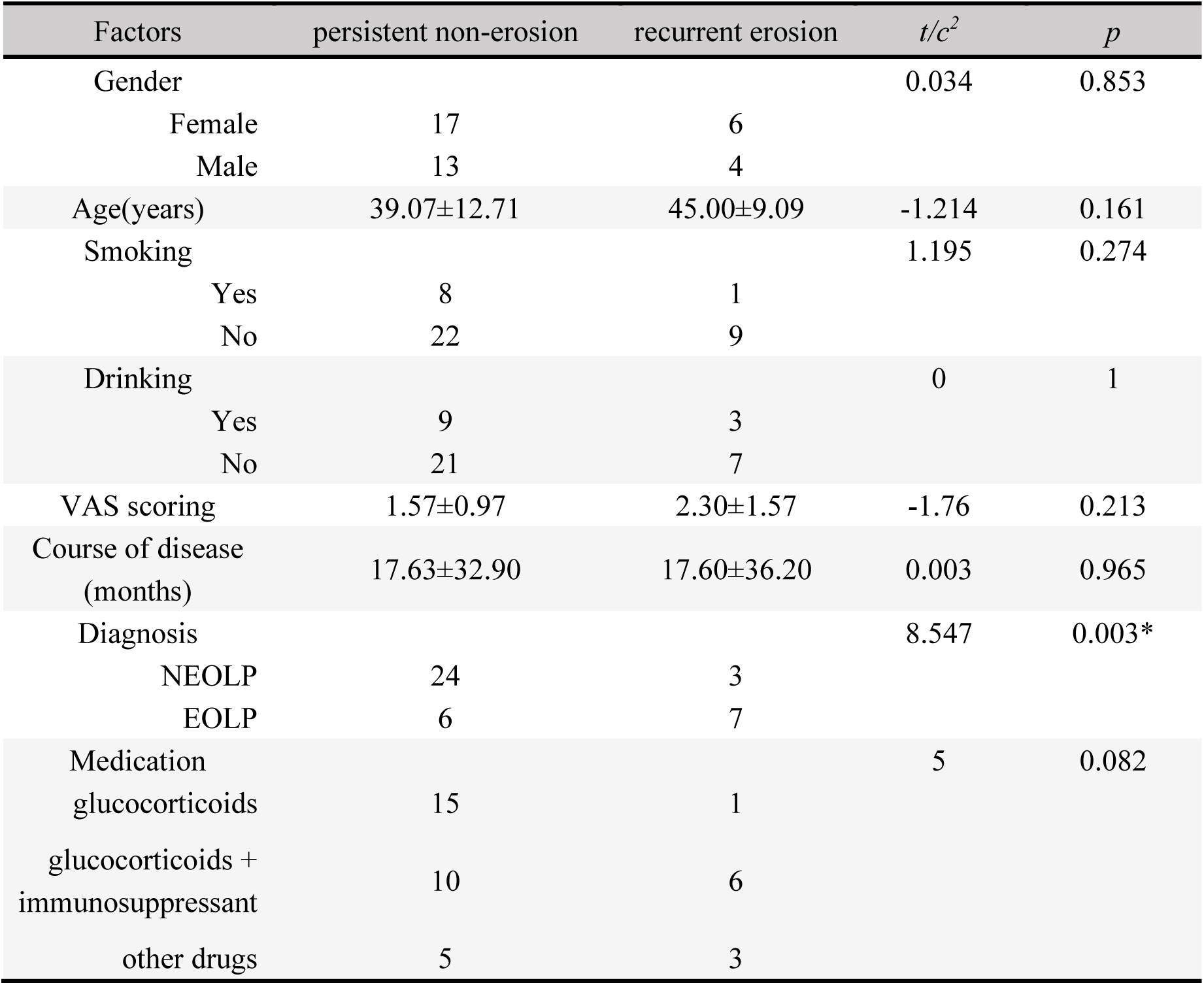
Analysis of clinical variables in different clinical outcomes of OLP.

To control the interference of confounding factors and avoid systematic bias, multivariate Logistic regression was used to further analyze patients with different clinical types and different clinical outcomes. The multivariate logistic model was used to perform regression analysis on the effects of CD8^+^ Trm-related factors on the clinical manifestations and clinical outcomes of OLP after adjusting for clinical factors such as age, gender, smoking, and drinking. We found that the core genes of CD8^+^ Trm cells are closely related to the clinical manifestations of OLP, and the expression differences of marker genes such as ITGA1, LITAF, SKIL, and cytokine-related genes such as IL17RA, IFNG41, IFNAR1, and TNFRSF6B were statistically significant between the two clinical types. The core genes of CD8^+^ Trm cells are also closely related to the erosion of OLP. Marker genes such as CD69, ITGA1, LITAF, SKIL, and cytokine-related genes such as IL17RA, IFNG41, IFNAR1, and GZMB are associated with two clinical outcomes of persistent non-erosion and recurrent erosion. The differences in expression were statistically significant and were significantly increased in the recurrent erosion group (Table 3).

#### CD8^+^ Trm cells may participate in the erosion of OLP by secreting active cytokines

Immunofluorescence experiments verified that there were CD8^+^ Trm cells in NEOLP and EOLP. The content of CD8^+^ Trm cells in EOLP was significantly increased than that in NEOLP. It can be seen from the location that CD8^+^ Trm cells are mostly close to the basal layer. In NEOLP, the basement membrane was basically intact, and sporadic T cells entered the epithelial layer, while in EOLP, CD8^+^ Trm cells accumulated more obviously under the erosive epithelium, and the number was obviously higher than in NEOLP, and the mucosal epithelial basement membrane was not clear in some places, more T cells were entering the mucosal epithelial layer, some of which were CD8^+^ Trm cells (Figures 4A-4D).

**Fig. 4.**
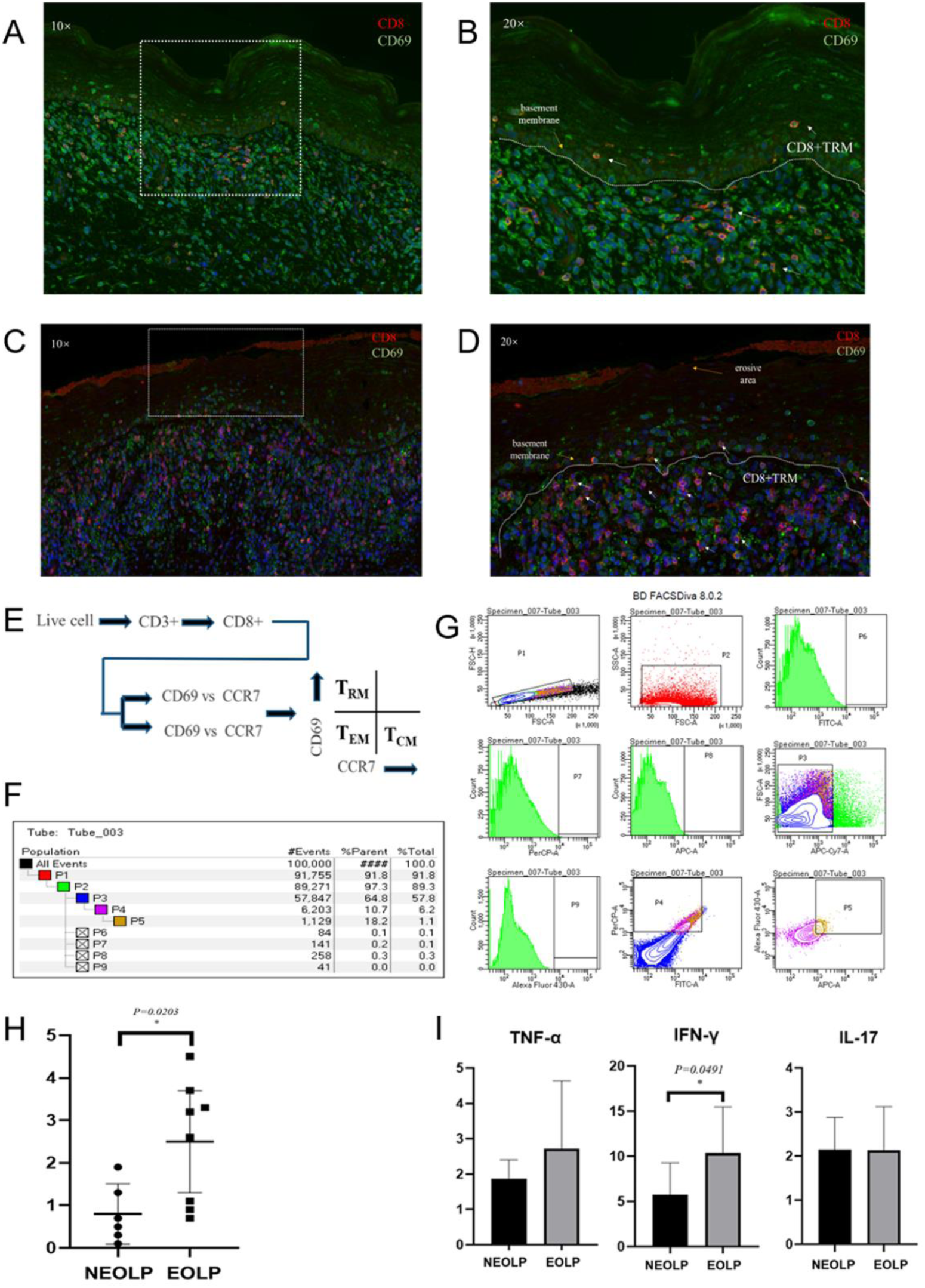
The increase of CD8+TRM in EOLP is closely related to the basement membrane and affects the clinical manifestations of OLP through its active secretion. (A-B) Immunofluorescence images of CD8+ TRM cells in NEOLP (The white dotted line is the basement membrane, and the red arrow points to the CD8+ TRM cells). (C-D)) Immunofluorescence images of CD8+ TRM cells in EOLP (The white dotted line is the basement membrane, and the red arrow points to the CD8+ TRM cells). (E) Schematic diagram of CD8+ TRM sorting by flow cytometry. (F-G) Image of sorting CD8+ TRM flow cytometry. (H) Comparison of the proportion of CD8+ TRM in NEOLP versus EOLP. (I) ELISA detection results of CD8+ TRM secreting TNF-α, IFN-γ and IL-17 in NEOLP versus EOLP.

According to the classic distinguishing and defining characteristics of memory T cell subsets, a CD8^+^ Trm sorting strategy was developed (Figure 4E). The OLP tissue cell suspension was incubated with antibodies for sorting, and it was found that the content of CD8^+^ Trm cells in EOLP was significantly higher than that in NEOLP (P < 0.05) (Figures 4F-4H). We further verified the previous results in scRNA-seq and ST and found that the difference in CD8^+^Trm content in different clinical types of OLP is strongly correlated with the clinical feature, which may be one of the reasons for the different clinical manifestations.

Then, we stimulated CD8^+^ Trm cells, which were sterile-sorted from EOLP and NEOLP tissues with phytohemagglutinin (PHA) and an enzyme-linked immunosorbent assay (ELISA) was performed to detect the supernatant of CD8^+^ Trm cells. The three inflammatory cytokines TNF-α, IFN-γ, and IL-17 produced by CD8^+^ Trm cells in EOLP were higher than in NEOLP, and the increase of INF-γ was statistically significant (Figure 4I). It also confirmed the previous experimental results and indicated that CD8^+^ Trm cells may affect the clinical manifestations of OLP through the secretion of the inflammatory factor IFN-γ.

## Discussion

Collectively, this study is the first to describe the presence, spatial distribution, and heterogeneity of CD8^+^ Trm cells in OLP tissues in different clinical manifestations, and to explore the regulatory mechanism of CD8^+^ Trm cells in OLP pathology. This study highlighted CD8^+^Trm cells involved in the pathological process of OLP and could promote erosion of OLP by secreting multiple cytokines.

We utilized scRNA-seq combined with spatial transcriptome to summarize the immune microenvironment of normal oral mucosa and OLP with different clinical subtypes. This study displayed that CD8^+^ Trm cells exist in OLP and normal tissues and the content of CD8^+^ Trm cells in normal mucosa, non-erosive OLP, and erosive OLP increased gradually. Compared with NEOLP, CD8^+^ Trm cells in EOLP were more extensive, especially in the epithelial deletion area. Pseudo-time analysis suggests that CD8^+^ Trm cells are one of the terminal states of T cells in OLP.

CD8^+^ Trm cells may affect the biological process of OLP by releasing cytokines such as TNF, IFN, IL-17, and the activity of CD8^+^ Trm cells was enhanced in EOLP. It is suggested that CD8^+^ Trm cells have an important relationship with the clinical type of OLP. Previous studies have found that CD8^+^ Trm cells can produce IFN-γ, a key cytokine in addition to viral infection(Cheuk et al., 2017; Guidotti et al., 1996). CD8^+^ Trms can also produce perforin and granzyme B upon IL-15 stimulation to mediate cytotoxic responses that respond to infections by enhancing local immune responses (Cheuk et al., 2017). In addition, IFN-γ upregulates the production of vascular cell adhesion molecule-1 (VCAM-1), which recruits TCM and B cells to the site of Trm cell localization (Ariotti et al., 2014).

Subsequently, we constructed an OLP clinical follow-up cohort and performed bulk RNA-seq of local lesions in patients. We found that compared with NEOLP, CD8^+^ Trm cells marker genes and related factors were significantly increased in EOLP. At the same time, after follow-up analysis, it was found that the CD8^+^ Trm marker genes and related immune factors in the recurrent erosion group were significantly increased than those in the persistent non-erosion group, which were consistent with the results of EOLP and also confirmed the consistency and accuracy of the results of EOLP. Multiple regression analyses based on OLP clinical cohort showed that multiple core genes of CD8^+^ Trm were closely related to OLP outcomes, which may be a significant factor leading to the deteriorative clinical outcome of OLP. It may cause repeated erosive lesions in OLP patients through local activation. Similarly, CD8+ Trm cells play a crucial part in the recurring aggravation of psoriasis; even after the psoriatic skin lesions have disappeared, CD8+ Trm cells can still be detected in the skin that seems normal. They have the potential to escalate local inflammation over time, trigger recurrent attacks at the same site, and generate a domino effect of inflammation (Owczarczyk Saczonek et al., 2020).

Moreover, immunofluorescence and flow cytometry experiments confirmed that the distribution of CD8^+^ Trm cells was closely related to the erosion of epithelial tissue, and its content in EOLP was significantly higher than that in NEOLP; An ELISA test confirmed that CD8^+^ Trm cells in EOLP tissue have enhanced ability to produce inflammatory cytokines such as IFN-γ compared with NEOLP, which may be an important promoting factor of epithelial erosion. Previous evidence suggests that CD8+ Trm cells protect the host against viruses by producing IFN-γ in lung tissue (Jiang et al., 2022a). Immune and inflammatory factors are considered to play an important role in the development of OLP, and cytokines IFN-γ may contribute greatly to the immune pathogenesis of OLP.

Currently, there are suggestions that one of the reasons of the recurrent episodes of many autoimmune diseases is the difficulty in totally eliminating local Trm cells(Jiang et al., 2012; Owczarczyk Saczonek et al., 2020). Immunosuppressive medications, local, or systemic corticosteroids are the main treatments for symptomatic OLP(Garcia-Pola et al., 2017; Raj et al., 2021). We discovered that there was no statistically significant difference between medication therapy and repeated degradation of OLP in our cohort analysis. This might be because CD8+ Trm is difficult for the present medications to entirely eradicate.

Therefore, through a series of studies, we confirmed that CD8^+^ Trm cells are increased in content and functionally active in EOLP, mainly distributed in the lamina propria close to the basement membrane, and released a variety of inflammatory factors, and clinical manifestations of OLP may be affected by the release of excess cytokines such as IFN-γ. Inflammatory mediators influence the clinical types and outcomes of OLP. CD8^+^ Trm cells may be a great important promoting factor for the recurrent erosion of OLP, which may become a potential immunotherapy target and provide new ideas for the treatment of OLP.

Our study has two limitations. Firstly, scRNA-seq was not performed on normal oral mucosal tissue to explore the role of CD8^+^ Trm cells in the pathogenesis of OLP。 Secondly, since there is currently no animal model for OLP, a cell co-culture model involving CD8^+^ Trm cells could provide more evidence in further studies. Despite these limitations, our findings support that CD8^+^ Trm cells play a considerable role in the disease process of OLP and a critical factor in the worsening of its clinical manifestations.

## EXPERIMENTAL MODEL AND SUBJECT DETAILS

### Human samples

All individuals provided written informed consent and this study was supported by the Ethics Committee of West China Hospital of Stomatology Sichuan University [WCHSIRB-2019-167]. According to the lesion with or without erosion in the biopsy, this study classified OLP as two groups, NEOLP and OLP.

### Cell isolation and processing

In the framework of the study, OLP tissues were either processed immediately for scRNA-seq or flow cytometry and cell sorting and stored at +80℃ refrigerator in freezing for bulk RNA-seq. All biopsied mucosa samples were immediately processed for scRNA-seq or flow cytometry and cell sorting. Samples were gently removed adipose tissue and minced with scissors in a sterile tissue culture dish.

Tissue fragments were then digested with enzyme mixture 500µl (whole-skin dissociation kit, Miltenyi Biotec) in gentleMACS C tubes (Miltenyi Biotec) which were incubated in a 37°C water bath for 3h under manual agitations every 15 min. Samples were mechanically separated with gentleMACS Dissociator (Miltenyi Biotec) for 1 min and then spun to collect.

Then move C tubes back to the water bath for another 20min. Next, enzymes were inactivated with 1ml precool DMES (Gibco Laboratories) at the end of the incubation. Samples were filtered through a 70-μm strainer (BD Bioscience) and collected by centrifuging in a table-top centrifuge at 400 × g at 4°C for 5 min. Samples were treated with 5ml ACK lysis buffer (Miltenyi Biotec) for 5 min and centrifuged at 400 × g at 4°C for 5 min. Remove the supernatant, and added 100ul dead cell removal kit (Miltenyi Biotec) for 15 min. Then added 1500ul into the cell suspension 3 times, and through the magnetic column (Miltenyi Biotec) after pipetting and mixing. Samples were centrifuged at 400 × g at 4°C for 5 min and resuspended in 200ul D-PBS.

### 10× Genomics library preparation and sequencing

The 10× Genomics Chromium System is a microfluidic platform based on Gel-bead in EMulsion (GEM) technology for generating real test datasets.

Gel beads containing barcode information were combined with a mixture of cells and enzymes and then encapsulated in microfluidic droplets to form GEMs.

The GEMs flowed into a reservoir and were collected, the gel beads were lysed to release the barcode sequences, the cDNA fragment was reverse transcribed, and samples were labeled. The gel beads were broken and the oil droplets were broken up and PCR amplification was performed using the cDNA as a template. The products of all GEMs are mixed and a standard sequencing library was constructed after using the Chromium Single Cell 5′ library or 3’ v2 library preparation kit according to the manufacturer’s protocol (10x Genomics). Finally, all sequencing experiments were conducted using Illumina NextSeq 500 in the Genomics Sequencing.

### Cell clustering and cell type annotation

The R package Seurat (v 4.0.1) was used to cluster the cells in the merged matrix. The first step was to filter out low-quality cells, which included cells with less than 500 transcripts, less than 100 genes, or cells with more than 10% of mitochondrial expression. And then normalized the data used the NormalizeData function. The canonical correlation analysis was performed using the normalized expression levels, the batch effect was corrected, and the data were integrated. Z-score normalization is performed on the integrated data, and principal component analysis (PCA) is performed using the normalized expression. The goal of PCA is to reduce the dimension of feature vectors by compressing the scale of the original data matrix, representing the most important features with the least dimension, and the new variable is a linear combination of the original variables, reflecting the comprehensive effect of the original variables. Dimensionality reduction through PCA reduced variables and finally clustered the cells.

### Cell-type subclustering and cell trajectory analysis

The prevalent cell types underwent subclustering. The subclusters were obtained using the identical functions as previously mentioned. We eliminated from further analysis subclusters that were solely defined by mitochondrial gene expression, a sign of low quality. By crossing over the canonical subtype signature genes with the marker genes for the subclusters, the subtypes were annotated. To identify the canonical pathways and putative upstream regulators, IPA was applied to the DEGs. Significant upstream regulators were those with activation z scores ≥2 or ≤2. Using the Add Module Score function on the genes activated by the targeted cytokine from bulk RNA-seq data, as previously stated, the module scores were generated.

In this study, Monocle 2 was used for cell trajectory analysis. First, all DEGs in cell subtypes (clusters) were screened; then dimensionality reduction was performed and then a minimum spanning tree was constructed; Finally, the best cell development or differentiation pseudo-time trajectory curve was fitted.

### Spatial Transcriptomics process

The Spatial Transcriptomics (ST) protocol was performed according to recommendations (10× Genomics). Fresh OLP tissues were put into a mold in powdered dry ice, wrapped with OCT, completely frozen and optimized to ideal form, and then sectioned while HE stained. The OCT-embedded tissues were cut to 10um thickness with a cryostat, then adhered to the chips, and frozen sectioned.

The chips with the tissue section were fixed with methanol, then stained with HE, and incubated with permeabilase on a PCR adapter to release the mRNA in the cells and bind to the corresponding capture probe. Validation of chips with brightfield imaging and fluorescence imaging. And then cDNA synthesis and sequencing library preparation using captured RNA as template.

Then, the prepared sequencing library is subjected to second-generation high-throughput short-read sequencing; finally, combined with the HE results, the expression of genes, the level of expression, and the spatial location information of these genes are determined.

### Spatial Transcriptomics analysis

Samples’ data were initially processed using the 10× official software Space Ranger (10×Genomics). Space Ranger displays the captured area organized in the chip through an image processing algorithm and distinguished the reads of each spot according to the spatial barcode information. The number of pair reads, the numbers of detected genes, and the number of UMIs in each spot were counted to evaluate the quality of the samples. The data were then normalized using sctransform to construct a regularized negative binomial model of gene expression to detect high variance features. After overall quality control, the proportion of mitochondrial genes in each sample is less than 0.12%, and the overall average is less than 0.08%, which meets the requirements of the 10x Genomics Visium platform.

After further dimensionality reduction, the gene expression of each spot was used to cluster the same type of spots to form spots clusters. Finally, through the labeling of characteristic genes, transcriptome visualization in the tissue space can be realized.

### Immunofluorescence staining

5um formalin-fixed mucosa lesion biopsy tissue sections were dewaxed in xylene and rehydrated with distilled water. Membrane rupture after proteinase K repair, then, the TDT enzyme, dUTP and buffer in the tunel kit were mixed at a ratio of 1:5:50, added to the tissue area, and then transferred to the wet box, and incubated a 37°C for 2 hours. After elution, the tissue surface was covered with 3% BSA and blocked at room temperature for 30 minutes. Then, incubation with the following primary monoclonal antibodies was performed: rabbit anti-human CD8, and mouse anti-human CD69, all at 1:200 dilution.

### Flow cytometry and cell sorting

Surface marker staining after resuspending the digested single cell suspension from the lesion of mucosa in 1-2ml of BD Pharmingen Stain Buffer (BSA) pre-chilled at 4°C. After rinsing twice with BSA, the cells were stored at 4°C in the dark, and flow cytometry was performed immediately. The BD cytometer was used and samples were sorted with fluorescence-activated cells, and the scheme of surface staining was as indicated in the text, using DAPI as the reactive dye.

### Enzyme-linked immunosorbent assay procedure

After sorting the CD8^+^ TRM cells, add 10ug/ml PHA to its 1640 medium. The supernatant was collected after culturing at 5% CO2 and 37°C for 72 hours, and then an ELISA assay was performed to detect TNF-α, IFN-γ, and IL-17 respectively.

Add deionized water to the corresponding reagents in the kit to prepare the detergent, diluent, and standard solution, prepare a standard dilution tube and prepare the color developer 15 minutes before use. Then, standard substances and experimental samples of different concentrations were added to the corresponding microwells, and 100ul was added to each microtiter well and incubated for 2h after sealing.

After washed of each well, added 200ul of enzyme-labeled detection antibody, then sealed the plate and incubated for 2 hours before washing, added 200ul of a diluted chromogenic substrate, sealed the plate and incubated for 30 minutes, and added 50ul of stop solution to each well. Measured the absorbance at 450nm with a microplate reader, set 540nm as the calibration wavelength, and measured the OD value of each well after zero-adjusting the blank control well. Finally, made a standard curve according to the concentration and OD value of the standard product, and then calculated the sample concentration according to the standard curve equation.

### Bulk-RNA sequencing procedure

Sample RNA was extracted by Cetyl Trimethyl Ammonium Bromide (CTAB) method ((Wang and Stegemann, 2010). And the extracted RNA was then tested for purity, concentration, and integrity. A number of library construction procedures, including end repair, end addition of A, ligation adapter addition, and fragment security screening, were carried out once mRNA is recovered. It was sequenced on the device following PCR amplification and purification.

### Clinical cohort and transcriptome data analysis

T-tests for continuous variables and chi-square tests for categorical variables were used to analyze the baseline data of the clinical cohort’s patients. T-tests were also used to determine whether one gene’s expression varied from that of another.

Multivariate logistic regression was used to analyze the effects of related genetic factors on the clinical outcome of OLP to account for confounding. Analysis based on a variety of variables. R software (Version 4.0.1) was used for the analysis and statistics of this portion of the statistical data, and all of the aforementioned analysis and test levels were set to 0.05.

Unreliable data was filtered and eliminated by the quality assessment of the original data after sequencing is complete, and downstream analysis was carried out following gene difference analysis. Correlation analysis, principal component analysis, sample cluster analysis, and weighted gene co-expression network analysis had next be performed.

## Supporting information

Including Figs. S1 to S2 and Table S1

## Data Availability

The data of this study, including scRNA-seq data, ST data, and bulk RNA-seq data are available in the Gene Expression Omnibus (GEO) database and the accession number is waiting to obtain.

## ACKNOWLEDGEMENTS

This work was supported by grants from the National Natural Science Foundation of China (No. 81730030 and No. 82001059). Thanks for the support of the department of oral medicine of West China Hospital of Stomatology.

## AUTHOR CONTRIBUTIONS

Conceptualization and study design: M.F.Q, Y.Z, H.X, and Q.M.C

Collection of clinical information and samples: M.F.Q, L.J

Performance of experiments: M.F.Q, Q.H.S

Data analysis: M.F.Q, J.K.P, D.Y, J.L

Figure preparation: M.F.Q, Q.H.S, J.X.D

The manuscript writing: M.F.Q, Q.H.S, H.X

Revision of the manuscript: all authors.

