## Supplementary material for "CD8+ tissue-resident memory T cells triggered the erosion of oral lichen planus by the cytokine network": Including Figs. S1 to S2 and Table S1

### Supplementary information

**This PDF file includes:**

Figs. S1 to S2
Table S1

**Fig. S1 The characteristics of NK/T cells.** (A) NK/T cells clustering in each OLP sample. (B) Heat map of differential genes in 0-10 cell subpopulations in NK/T cells. (C) Heat map of significant differentially expressed genes (DEGs) in NK/T cells subsets.

**Fig. S2 CD8+ TRM in OLP patients have different transcriptomic landscapes in different clinical presentations and outcomes.** (A-B) CD8+ TRM-related cytokine (product and/or receptor) marker genes in EOLP/NEOLP. (C-D) CD8+ TRM-related cytokine (product and/or receptor) marker genes in RE/PNE.

**Table S1 Number of Spots after quantification.**

**Fig. S1 The characteristics of NK/T cells.** (A) NK/T cells clustering in each OLP sample. (B) Heat map of differential genes in 0-10 cell subpopulations in NK/T cells. (C) Heat map of significant differentially expressed genes (DEGs) in NK/T cells subsets.

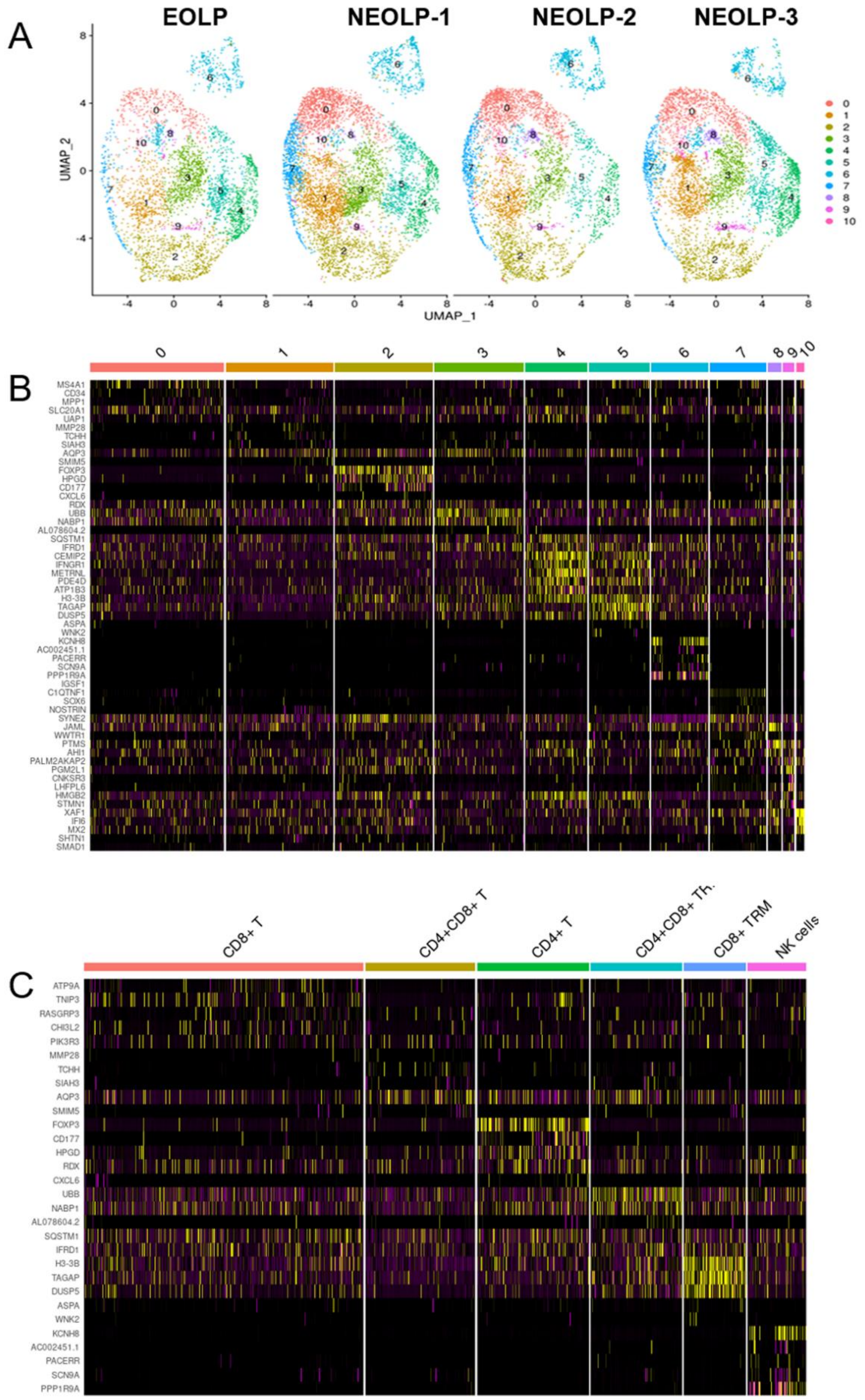

**Fig. S2 CD8+ TRM in OLP patients have different transcriptomic landscapes in different clinical presentations and outcomes.** (A-B) CD8+ TRM-related cytokine (product and/or receptor) marker genes in EOLP/NEOLP. (C-D) CD8+ TRM-related cytokine (product and/or receptor) marker genes in RE/PNE.

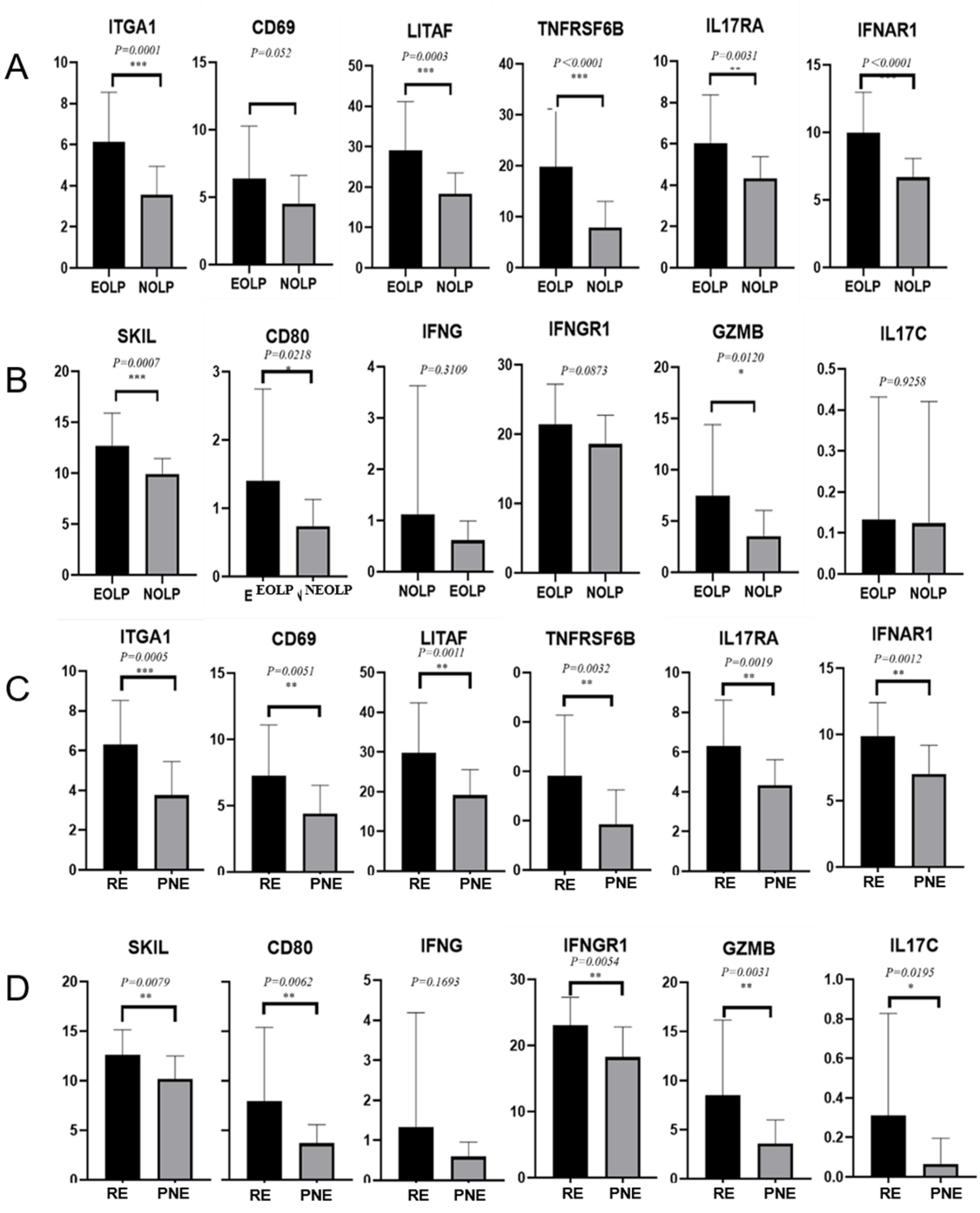

**Table S1 Number of Spots after quantification.**

| Samples | Mean_nUMI | Mean_nGene | Total spots |
|---|---|---|---|
| EOLP | 18975.35736 | 3223.513514 | 666 |
| NOLP-1 | 8532.503378 | 2869.192568 | 296 |
| NOLP-2 | 11239.82887 | 3119.470238 | 672 |
| NOLP-3 | 16840.56891 | 3322.493264 | 965 |
| Normal-1 | 14328.00909 | 3442.531818 | 440 |
| Normal-2 | 15171.08961 | 3883.52509 | 558 |
